# Association of 24-Hour Urinary Sodium-to-Potassium Ratio with Deep White Matter Lesions in Community-Dwelling Older Adults

**DOI:** 10.64898/2026.06.29.26356891

**Authors:** Kenji Fukuda, Hiroshi Yao, Masahiro Kamouchi, Toru Nabika, Mari Mori, Hideki Mori, Yasushi Okada, Yukio Yamori, Tetsuro Ago

## Abstract

**Background:** The urinary sodium-to-potassium (Na/K) ratio is an integrated biomarker associated with cardiovascular risk. However, its association with deep white matter lesions (DWMLs), a manifestation of cerebral small vessel disease (CSVD), remains unclear. We investigated the associations of the urinary Na/K ratio and albuminuria with DWMLs.

**Methods:** We conducted a cross-sectional study of 296 Japanese adults (mean age, 68.7 years). Brain magnetic resonance imaging was used to assess DWMLs using the Fazekas scale, and lesions were classified as absent (grade 0) or present (grades 1–3). The urinary Na/K ratio and albumin excretion were measured using 24-h urine collections. Multivariable logistic regression models examined the associations between urinary biomarkers and DWMLs.

**Results:** DWMLs were present in 119 (40.2%) participants. A higher urinary Na/K ratio was independently associated with DWMLs (odds ratio per 1–standard deviation increase, 1.44; 95% confidence interval, 1.09–1.90; P=0.010). Participants in the highest quartile had greater odds of DWMLs than those in the lowest quartile (odds ratio, 2.48; 95% confidence interval, 1.16–5.29; P=0.019). Urinary potassium excretion was inversely associated with DWMLs, whereas urinary sodium excretion alone showed no significant association. Findings were consistent across sensitivity and subgroup analyses.

**Conclusions:** A higher 24-h urinary Na/K ratio was independently associated with DWMLs in older adults. This association appeared to be driven primarily by lower urinary potassium excretion rather than higher sodium excretion alone. The urinary Na/K ratio may serve as a simple, noninvasive marker of CSVD.

**Clinical Perspective:** *What Is New?:*

- In this community-based study of older adults, a higher 24-h urinary sodium-to-potassium ratio was independently associated with the presence of deep white matter lesions on brain magnetic resonance imaging.
- The association seemed to be driven primarily by lower urinary potassium excretion rather than by higher sodium excretion.

*What Are the Clinical Implications?:*

- The urinary sodium-to-potassium ratio may serve as a simple, noninvasive biomarker of cerebral small vessel disease.
- Improvement in dietary sodium-to-potassium balance, particularly through increased potassium intake, may contribute to the preservation of cerebral microvascular health.
- Measurement of the urinary sodium-to-potassium ratio may provide additional information for cerebrovascular risk assessment.

## Introduction

White matter lesions (WMLs), a hallmark of cerebral small vessel disease (CSVD), are frequently observed in older adults and are associated with increased risks of cognitive decline, stroke, and functional impairment in daily life.^1,2^ Chronic ischemia and vascular inflammation represent the primary mechanisms underlying the WML development, whereas hypertension, diabetes, smoking, and dyslipidemia are recognized as key contributing factors.^3,4^ Despite their clinical importance, the influence of modifiable dietary factors on WMLs remains insufficiently investigated.

The urinary sodium-to-potassium (Na/K) ratio reflects the balance between sodium and potassium intake and serves as a practical biomarker of habitual dietary electrolyte consumption. Because most sodium and a substantial proportion of potassium are excreted in urine, the urinary Na/K ratio provides a useful measure of long-term dietary patterns and salt-related dietary habits.^5–9^

Albuminuria, a marker of systemic endothelial dysfunction, has been associated with CSVD and may serve as an early indicator of cerebrovascular pathology.^10,11^ However, evidence regarding the association between the urinary Na/K ratio and WMLs, particularly deep white matter lesions (DWMLs), remains limited. Furthermore, few studies have simultaneously evaluated the urinary Na/K ratio and albuminuria using 24-h urine collections. Clarifying these associations may improve understanding of potentially modifiable risk factors and facilitate the identification of individuals at increased risk of CSVD. To our knowledge, few studies have simultaneously investigated the urinary Na/K ratio and albuminuria in relation to DWMLs using 24-h urine collections.

In this study, we aimed to investigate the associations of the urinary Na/K ratio and albuminuria with the presence of DWMLs in community-dwelling Japanese adults. The potential utility of these urinary biomarkers for identifying DWMLs, a manifestation of CSVD, was evaluated using 24-h urine collections and brain magnetic resonance imaging (MRI).

## Methods

### Participants and Protocol Approval

A cross-sectional study was conducted in two neighboring rural communities in Saga, Japan, between 2015 and 2019. A total of 475 community-dwelling adults (aged 39–89 years; 90.5% aged ≥60) volunteered to participate. All participants were independent in activities of daily living, free of dementia, and underwent brain MRI. Among them, 381 (80.2%) provided 24-h urine samples using a validated aliquot device that collected 1/40 of each void. After excluding 29 individuals who did not meet the predefined MRI eligibility criteria (e.g., history of stroke, brain tumor, or claustrophobia) and 56 individuals with incomplete or unreliable urine samples, 296 participants were included in the final analysis. The Institutional Review Board of the National Hospital Organization Hizen Psychiatric Center (approval numbers: 15-1 and 24-4) approved the study, and all participants provided written informed consent. All procedures were conducted in accordance with relevant guidelines and regulations.

### Clinical Assessments

All participants underwent a structured clinical interview and general hematological and biochemical testing. Simultaneous office blood pressure (BP) measurements were obtained in the seated position from both arms using an automated sphygmomanometer (Omron HEM-1020; Omron, Japan), and the mean of the two readings was used to characterize clinical status. Hypertension was diagnosed based on a documented medical history with repeated BP measurements ≥140/90 mmHg or current use of antihypertensive medication rather than the single office BP measurement obtained during the study visit.

Vascular risk factors were defined as follows. Metabolic syndrome was defined as central obesity plus at least two of the following conditions: elevated BP (≥130/85 mmHg), elevated fasting glucose (≥110 mg/dL), or dyslipidemia (triglycerides ≥150 mg/dL and/or HDL cholesterol <40 mg/dL), based on established criteria. Diabetes mellitus was defined as a fasting plasma glucose level ≥126 mg/dL, HbA1c ≥6.5%, or a previous diabetes diagnosis. Hyperlipidemia was defined as a total serum cholesterol level ≥220 mg/dL or the use of lipid-lowering medications.

Estimated glomerular filtration rate (eGFR) was calculated using the Japanese-modified MDRD equation. Current smokers were defined as individuals who smoked ≥10 cigarettes per day, whereas former smokers were classified as nonsmokers. Alcohol use was defined as consumption of ≥4 alcoholic drinks (40 g ethanol) per week.

### Measurement of 24-h Urine Indices

All measurements were performed between late September and early December. An aliquot cup was used for 24-h urine collection, and one-fortieth of the voided urine volume was sampled at each urination. Participants practiced using the aliquot cup with water before the actual urine collection and received instructions to collect 24-h urine accurately. After discarding the first morning urine sample, participants collected all urine samples during the subsequent 24-h period. Urine samples were frozen at −80 °C until analysis.

Urinary sodium (Na) and potassium (K) concentrations were determined using the electrode method, and creatinine levels were measured using the enzymatic method. The 24-h urinary albumin level was determined using the turbidimetric immunoassay method. For descriptive analyses, urinary sodium and potassium excretion were expressed as sodium chloride (NaCl, g/day) and potassium (g/day), respectively. For regression analyses, sodium and potassium excretion values were converted to mmol/day to calculate the urinary Na/K ratio, which is unitless.

### MRI and Assessment of WMLs

Brain imaging was performed using a 1.5-T MRI scanner (Achieva; Philips Healthcare, Best, the Netherlands). A combination of T1-weighted, T2-weighted, and fluid-attenuated inversion recovery (FLAIR) sequences was used to detect DWMLs. DWMLs were defined as areas that were isointense to normal parenchyma on T1-weighted images and hyperintense on both T2-weighted and FLAIR images. Lesions were graded using the validated Fazekas scale as follows: grade 0, absent; grade 1, punctate foci; grade 2, beginning confluence of foci; and grade 3, large confluent areas.^1^ Two trained researchers (H.Y. and A.U.) who were blinded to the clinical data independently evaluated all scans. For statistical analyses, DWMLs were dichotomized as absent (Fazekas grade 0) or present (Fazekas grades 1–3).

### Statistical Analysis

Continuous variables are expressed as mean ± standard deviation (SD), and categorical variables as frequency (percentage). Between-group comparisons were performed using the unpaired t test for continuous variables and the χ^2^ test for categorical variables. Statistical significance was set at P < 0.05, unless otherwise specified. Univariate logistic regression analyses were conducted to identify variables associated with the presence of DWMLs. Variables with statistically significant associations in the univariate analyses were included in the multivariable models. Age and sex were included in all models regardless of statistical significance because of their clinical relevance. Three sequential multivariable logistic regression models were constructed to evaluate the independent associations between urinary parameters and DWMLs. Model 1 included age, sex, and conventional vascular risk factors identified in the univariate analyses. Model 2 additionally included urinary albumin excretion, and model 3 further incorporated the urinary sodium-to-potassium (Na/K) ratio. Additional analyses focusing on the urinary Na/K ratio were performed to further characterize its association with DWMLs. These analyses included dose–response analyses based on quartiles of the Na/K ratio and continuous analyses per 1–SD increase in urinary sodium, potassium, and the Na/K ratio. Sensitivity analyses were conducted to assess the robustness of the primary findings, including analyses excluding hypertension from the covariates. Subgroup analyses stratified by clinically relevant variables were also performed, and multiplicative interaction terms were used to evaluate potential effect modification. All statistical analyses were conducted using IBM SPSS Statistics for Macintosh (version 23.0; IBM Corp., Armonk, NY, USA).

## Results

### Baseline Characteristics

A total of 296 community-dwelling adults (108 men and 188 women; mean age, 68.7 years) were included. DWMLs were present in 119 participants (40.2%) and absent in 177 participants (59.8%). According to the Fazekas scale, the majority of participants had absent or mild DWMLs: 59.8% were classified as grade 0, 28.7% as grade 1, 10.8% as grade 2, and only 0.7% as grade 3. Table 1 shows the summary of the baseline characteristics by DWML status. Participants with DWMLs were significantly older and had a higher prevalence of hypertension, higher systolic BP, more frequent use of antihypertensive medications, lower mean eGFR, a higher prevalence of reduced eGFR (<60 mL/min/1.73 m^2^), and higher geometric mean levels of 24-h urinary albumin excretion and the urinary Na/K ratio.

**Table 1.** Baseline Characteristics of Participants According to the Presence of DWMLs.

|  | DWMLs (–)<br>(n=177) | DWMLs (+)<br>(n=119) | P-value |
| --- | --- | --- | --- |
| Age, years, mean±SD | 67.0±7.6 | 71.3±5.8 | <0.001 |
| Female sex, n (%) | 107 (60.5) | 81 (68.1) | 0.22 |
| Body mass index, kg/m<sup>2</sup>, mean±SD | 23.3±3.4 | 23.7±3.3 | 0.36 |
| Hypertension, n (%) | 55 (31.1) | 70 (58.8) | <0.001 |
| Antihypertensive agents, n (%) | 46 (26.0) | 61 (51.3) | <0.001 |
| SBP, mmHg, mean±SD | 140.2±22.0 | 149.1±19.9 | <0.001 |
| DBP, mmHg, mean±SD | 76.8±11.7 | 78.9±9.9 | 0.11 |
| Metabolic syndrome, n (%) | 11 (6.2) | 12 (10.1) | 0.27 |
| Diabetes mellitus, n (%) | 22 (12.4) | 18 (15.1) | 0.60 |
| Dyslipidemia, n (%) | 67 (37.9) | 44 (37.0) | 0.90 |
| Smoking, n (%) | 7 (4.0) | 5 (4.2) | 0.99 |
| Alcohol, n (%) | 58 (32.8) | 38 (31.9) | 0.90 |
| History of coronary heart disease, n (%) | 3 (1.7) | 7 (5.9) | 0.10 |
| Hemoglobin A1c, %, mean±SD | 5.84±0.51 | 5.95±0.60 | 0.09 |
| LDL cholesterol, mg/dL, mean±SD | 121.7±29.0 | 115.0±28.0 | 0.05 |
| HDL cholesterol, mg/dL, mean±SD | 68.7±15.8 | 69.0±18.7 | 0.66 |
| Triglyceride, mg/dL, mean±SD | 127.8±62.8 | 136.6±87.0 | 0.31 |
| eGFR, mL/min/1.73 m<sup>2</sup>, mean ±SD | 71.0±12.2 | 66.1±13.1 | 0.001 |
| eGFR < 60 mL/min/1.73 m<sup>2</sup>, n (%) | 25 (14.1) | 35 (29.4) | 0.002 |
| 24-hour urine indices |  |  |  |
| Urinary albumin<sup>a</sup>, mg/day, geometric mean±SD | 0.94±0.34 | 1.09±0.45 | 0.001 |
| Urinary sodium (NaCl), g/day, mean±SD | 9.14±3.33 | 9.55±3.38 | 0.30 |
| Urinary potassium (K), g/day, mean±SD | 1.85±0.66 | 1.75±0.66 | 0.18 |
| Urinary Na/K ratio | 3.53±1.36 | 3.94±1.63 | 0.02 |
| Urinary Mg, mg/day, mean±SD | 74.1±31.5 | 70.7±31.3 | 0.36 |

Values are presented as mean ± standard deviation (SD) or number (%). <sup>a</sup>Urinary albumin values were log-transformed to approximate normality, and geometric means were reported. Abbreviations: DWMLs, deep white matter lesions; BMI, body mass index; SBP, systolic blood pressure; DBP, diastolic blood pressure; eGFR, estimated glomerular filtration rate; Na/K ratio, sodium-to-potassium ratio.

### Primary and Dose–Response Analyses

In the multivariable logistic regression analyses (Table 2), increasing age (odds ratio [OR] per 10 years = 2.33; 95% confidence interval [CI], 1.51–3.60; P < 0.001) and hypertension (OR = 2.92; 95% CI, 1.75–4.87; P < 0.001) were significantly associated with the presence of DWMLs in model 1, whereas eGFR was not significantly associated with DWMLs. After additional adjustment for urinary albumin in model 2, urinary albumin excretion showed a borderline association with DWMLs (OR = 1.83; 95% CI, 0.90–3.74; P = 0.097). Model 3 further included the urinary sodium-to-potassium (Na/K) ratio. A higher urinary Na/K ratio was independently associated with DWMLs (OR per 1–SD increase = 1.44; 95% CI, 1.09–1.90; P = 0.010), whereas the association between urinary albumin and DWMLs was attenuated and no longer statistically significant (OR = 1.74; 95% CI, 0.84–3.62; P = 0.135).

**Table 2.** Multivariable Logistic Regression Models for the Association of Urinary Albumin and the Na/K Ratio with DWMLs.

|  | Model 1 |  |  | Model 2 |  |  | Model 3 |  |  |
| --- | --- | --- | --- | --- | --- | --- | --- | --- | --- |
|  | OR | 95% CI | P-value | OR | 95% CI | P-value | OR | 95% CI | P-value |
| Age / 10 years | 2.33 | 1.51–3.60 | <0.001 | 2.34 | 1.51–3.63 | <0.001 | 2.47 | 1.58–3.87 | <0.001 |
| Female sex | 1.49 | 0.88–2.54 | 0.138 | 1.45 | 0.85–2.47 | 0.176 | 1.58 | 0.91–2.73 | 0.102 |
| Hypertension | 2.92 | 1.75–4.87 | <0.001 | 2.52 | 1.47–4.31 | 0.001 | 2.39 | 1.39–4.11 | 0.002 |
| eGFR | 0.99 | 0.97–1.01 | 0.290 | 0.99 | 0.97–1.01 | 0.337 | 0.99 | 0.96–1.01 | 0.191 |
| Urinary albumin <sup>a</sup> |  |  |  | 1.83 | 0.90–3.74 | 0.097 | 1.74 | 0.84–3.62 | 0.135 |
| Urinary Na/K ratio (per 1–SD) |  |  |  |  |  |  | 1.44 | 1.09–1.90 | 0.010 |

Model 1 was adjusted for age, sex, hypertension, and estimated glomerular filtration rate (eGFR).
Model 2 was additionally adjusted for 24-h urinary albumin.
Model 3 was further adjusted for the urinary sodium-to-potassium (Na/K) ratio.
<sup>a</sup> Urinary albumin was log-transformed to approximate normality.
Odds ratios (ORs) are presented per 1–standard deviation (SD) increase for continuous variables.
Abbreviations: DWMLs, deep white matter lesions; OR, odds ratio; CI, confidence interval; eGFR, estimated glomerular filtration rate; Na/K ratio, sodium-to-potassium ratio; SD, standard deviation.

In the quartile analyses of the urinary Na/K ratio (Table 3), participants in the highest quartile (Q4: 4.44–9.67) had significantly higher odds of DWMLs than those in the lowest quartile (Q1: 1.09–2.74) after multivariable adjustment (OR = 2.48; 95% CI, 1.16–5.29; P = 0.019). A significant linear trend was observed across quartiles (P for trend = 0.033).

**Table 3.** Association Between the Urinary Sodium-to-Potassium Ratio and DWMLs Across Quartiles.

|  | Number of<br>DWMLs + (%) | Model 1 |  |  | Model 2 |  |  |
| --- | --- | --- | --- | --- | --- | --- | --- |
|  |  | OR | 95% CI | P-value | OR | 95% CI | P-value |
| Q1 (1.09-2.74), n=75 | 25 (33.3) | 1.00 | reference |  | 1.00 | reference |  |
| Q2 (2.76-3.39), n=73 | 30 (41.1) | 1.51 | 0.74-3.06 | 0.256 | 1.84 | 0.87-3.93 | 0.113 |
| Q3 (3.42-4.41), n=74 | 27 (36.5) | 1.45 | 0.71-2.97 | 0.309 | 1.63 | 0.77-3.46 | 0.201 |
| Q4 (4.44-9.67), n=74 | 37 (50.0) | 2.63 | 1.29-5.38 | 0.008 | 2.48 | 1.16-5.29 | 0.019 |
| *P for trend (quartiles)* |  |  |  | 0.013 |  |  | 0.033 |

Odds ratios (ORs) and 95% confidence intervals (CIs) are shown for each quartile of the urinary Na/K ratio, with the lowest quartile as the reference.
Model 1 was adjusted for age and sex.
Model 2 was additionally adjusted for hypertension, estimated glomerular filtration rate (eGFR), and 24-h urinary albumin.
P for linear trend was calculated by modeling quartiles of the urinary Na/K ratio as an ordinal variable.
Abbreviations: DWMLs, deep white matter lesions; OR, odds ratio; CI, confidence interval; Na/K ratio, sodium-to-potassium ratio; Q, quartile; eGFR, estimated glomerular filtration rate.

### Continuous Associations of Urinary Electrolytes

Figure 1 shows the adjusted odds ratios per 1–SD increase in urinary electrolyte parameters. In this continuous-variable analysis, the urinary Na/K ratio demonstrated a clear positive association with DWMLs (OR per 1–SD increase = 1.44; 95% CI, 1.09–1.90; P = 0.010). Urinary potassium excretion was inversely associated with DWMLs (OR = 0.76; 95% CI, 0.59–0.99; P = 0.042), whereas urinary sodium excretion alone was not significantly associated (OR = 1.12; 95% CI, 0.86–1.47; P = 0.404).

**Figure 1.**
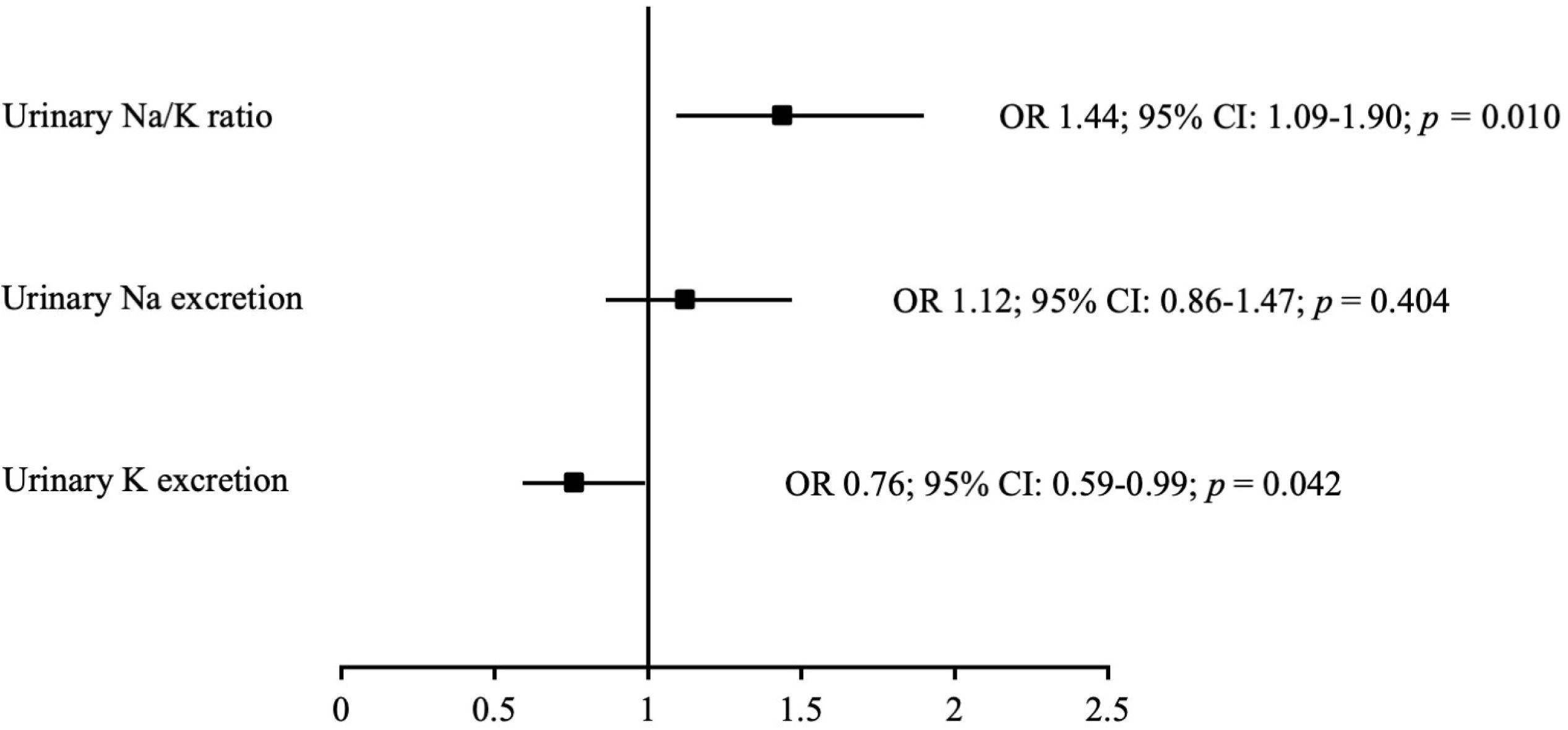
Adjusted associations of 24-h urinary electrolyte indices with deep white matter lesions. Adjusted odds ratios (ORs) and 95% confidence intervals (CIs) for the presence of deep white matter lesions (DWMLs) per 1-standard deviation (SD) increase in urinary sodium (Na), potassium (K), and sodium-to-potassium (Na/K) ratio. Models were adjusted for age, sex, hypertension, estimated glomerular filtration rate (eGFR), and 24-h urinary albumin excretion. Abbreviations: DWMLs, deep white matter lesions; OR, odds ratio; CI, confidence interval; SD, standard deviation; Na, sodium; K, potassium; Na/K ratio, sodium-to-potassium ratio; eGFR, estimated glomerular filtration rate.

### Robustness and Sensitivity Analyses

Sensitivity analyses excluding hypertension from the covariates demonstrated that urinary albumin levels showed significant association with DWMLs, whereas the urinary Na/K ratio remained independently associated with DWMLs (Supplementary Table S1).

### Subgroup Analyses

The association between the urinary Na/K ratio and DWMLs was consistent across subgroups defined by age (<70 vs. ≥70 years), sex, hypertension status, renal function (eGFR <60 vs. ≥60 mL/min/1.73 m^2^), and albuminuria status (<30 vs. ≥30 mg/day). No significant effect modification was observed in any subgroup (all P for interaction > 0.05) (Figure 2, Supplementary Table S2).

**Figure 2.**
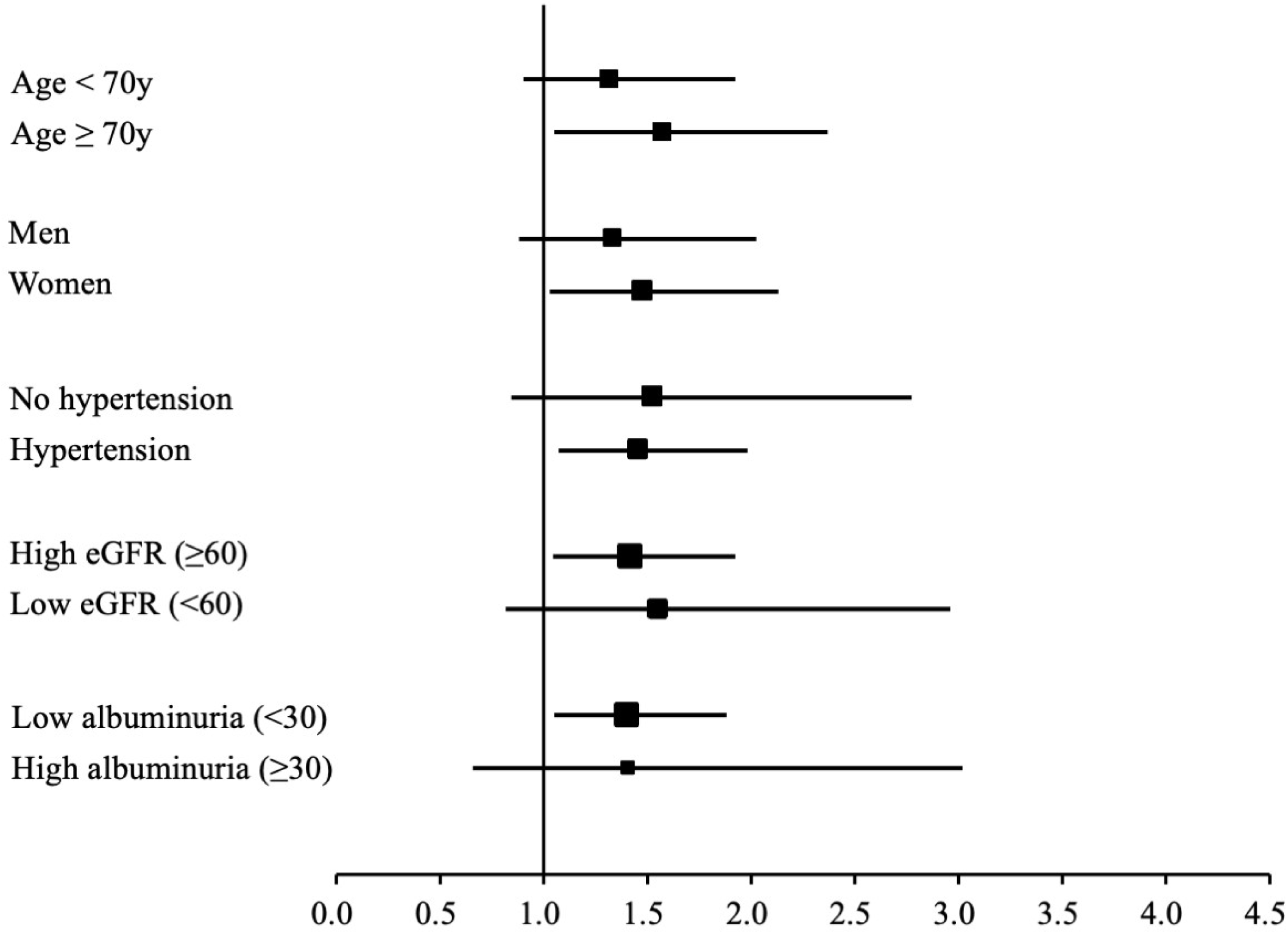
Consistency of the association between urinary Na/K ratio and DWMLs across clinical subgroups. Forest plot showing adjusted odds ratios (ORs) and 95% confidence intervals (CIs) for the association between the urinary sodium-to-potassium (Na/K) ratio and the presence of deep white matter lesions (DWMLs) across predefined subgroups. Subgroups were defined according to age (<70 vs. ≥70 years), sex, hypertension status, renal function (eGFR <60 vs. ≥60 mL/min/1.73 m^2^), and albuminuria (<30 vs. ≥30 mg/day). No statistically significant interactions were observed between the urinary Na/K ratio and any subgroup (all interaction P values >0.05). Subgroup-specific participant numbers and event frequencies are provided in Supplementary Table S2. Abbreviations: DWMLs, deep white matter lesions; OR, odds ratio; CI, confidence interval; Na/K ratio, sodium-to-potassium ratio; eGFR, estimated glomerular filtration rate.

## Discussion

To our knowledge, this community-based study is the first to demonstrate a statistically significant and independent association between the 24-h urinary Na/K ratio and DWMLs detected by brain MRI. In this cross-sectional analysis of 296 older adults, a higher urinary Na/K ratio was consistently associated with the presence of DWMLs across multivariable-adjusted logistic regression models, quartile-based dose–response analyses, and standardized continuous variable models. This association remained significant after adjustment for hypertension and other vascular risk factors. By contrast, 24-h urinary albumin levels showed a weaker, nonsignificant association after BP adjustment, whereas eGFR was not independently associated with DWMLs. Taken together, these findings suggest that the urinary Na/K ratio may serve as a novel and easily measurable biomarker of dietary electrolyte balance relevant to cerebral small vessel health.

The findings further indicate that the association between the urinary Na/K ratio and DWMLs was primarily driven by urinary potassium excretion. Urinary potassium excretion was inversely associated with lesion presence, whereas urinary sodium excretion alone showed no significant association. Because urinary potassium excretion is generally considered a surrogate marker of habitual dietary potassium intake, the observed association may reflect the vascular benefits of potassium-rich dietary patterns rather than potassium excretion itself. In the continuous variable analysis, each 1-SD increase in urinary potassium was associated with 24% lower odds of DWMLs. This finding is consistent with evidence from prospective cohort studies,^12^ meta-analyses and observational data,^13^ and more recent reviews and intervention studies demonstrating improvements in endothelial function, reductions in arterial stiffness, and modulation of the renin–angiotensin–aldosterone system.^14,15^ These observations align with previous reports showing that the urinary Na/K ratio more sensitively captures the combined and opposing vascular effects of sodium and potassium than either electrolyte alone.^9,16–18^ Consequently, the urinary Na/K ratio may represent a clinically practical index of dietary quality and salt sensitivity.

Albuminuria has been widely recognized as a marker of systemic microvascular injury and has been associated with CSVD in meta-analyses and epidemiological studies.^10,11,19^ However, in the present analysis, the association between albuminuria and DWMLs became non-significant after adjustment for hypertension. This attenuation supports the hypothesis that albuminuria reflects microvascular injury largely mediated by elevated BP. Supplementary analyses excluding participants with hypertension demonstrated a statistically significant association between albuminuria and DWMLs, indicating that the effect of albuminuria on DWMLs is strongly BP dependent. This finding is consistent with the “strain vessel” hypothesis, which proposes that glomerular and deep cerebral perforating arterioles, owing to their anatomical susceptibility to high intraluminal pressures, are vulnerable to pressure-mediated injury.^10^

The association between the urinary Na/K ratio and DWMLs remained significant regardless of whether hypertension was included as a covariate. This persistence suggests that the urinary Na/K ratio may capture factors beyond the effects of BP, including dietary sodium load, impaired sodium excretion, salt sensitivity, or cumulative dietary patterns that influence cerebral microvascular integrity.^20^

The present cohort predominantly comprised participants with absent or mild DWMLs, whereas advanced confluent DWMLs were relatively uncommon. Within a population enriched with early-stage WMLs, factors associated with functional or potentially reversible microvascular alterations may be more readily detected. This distribution may partly explain why the association with the urinary Na/K ratio was observed mainly with the presence of DWMLs rather than with lesion severity.

Given the simplicity and low cost of assessing the Na/K ratio from 24-h urine collections, and potentially from spot urine samples when 24-h collection is not feasible, this biomarker could be readily integrated into routine health screenings and public health programs. In Japan, where dietary sodium intake remains high and potassium intake remains suboptimal, these findings support existing national guidelines that advocate sodium reduction and potassium enrichment as a dual dietary strategy to reduce cardiovascular and cerebrovascular risk. Such measures may contribute to cerebral microvascular health by mitigating CSVD-related brain changes and preserving cognitive function.

This study has several strengths, including the use of the gold-standard 24-h urine collection method to assess sodium, potassium, and albumin excretion, as well as the MRI-based Fazekas scale for standardized DWML assessment.^1,2^ The combination of these precise measurements in a community-based cohort enhances the validity of the findings and supports their generalizability to similar populations. However, several limitations should be acknowledged. First, the cross-sectional design precludes causal inference. Second, 24-h urine collection, although highly accurate, may introduce selection bias by preferentially including more health-conscious individuals.^21,22^ Third, urinary sodium and potassium excretion were assessed from a single 24-hour urine collection, which may not fully capture day-to-day variation in dietary intake and sodium–potassium balance. Fourth, dichotomization of DWML grades may have reduced sensitivity to differences in mild lesion severity.^23^ Fifth, unmeasured confounding factors, including physical activity, dietary patterns beyond sodium and potassium intake, and genetic predisposition, may have influenced the observed associations. Finally, recruitment from rural Japanese communities may limit the applicability of the findings to other populations and settings.

In conclusion, the 24-h urinary Na/K ratio was independently associated with DWMLs in older adults. This association appeared to be influenced mainly by potassium excretion and persisted independently of BP. Albuminuria also demonstrated a BP-dependent association, consistent with the shared microvascular vulnerability of the kidney and brain. These findings highlight the potential importance of dietary sodium-to-potassium balance in maintaining cerebral small-vessel health. Longitudinal and mechanistic studies are warranted to confirm causality, elucidate the underlying biological pathways, and evaluate the utility of the urinary Na/K ratio as a target for screening and intervention in diverse populations.

## Data Availability

The datasets generated and/or analyzed during the current study are available from the corresponding author upon reasonable request.

## Acknowledgements

We thank Sachiko Kawasaki-Tsuchida for the technical support and Yoko Bando and Yuko Araki for the assistance with the statistical analyses. We thank Akira Uchino for reviewing all the MRI scans. We are also grateful to the staff of the National Hospital Organization Hizen Psychiatric Center for their long-standing cooperation in the development and maintenance of the Sefuri-Yoshinogari Study Database, which provided the foundation for this research. We express our sincere gratitude to Eno Honda for the technical support and assistance with manuscript preparation and submission.

## Author Contributions

All authors significantly contributed to this research. K.F. and H.Y. conceived the study, designed the study protocol, and supervised the project. H.Y., M.K., and Y.O. contributed to the acquisition and interpretation of the data; H.Y. was a primary investigator of the Sefuri-Yoshinogari Study at the National Hospital Organization Hizen Psychiatric Center from 1997 to 2021. T.N. contributed to the methodology, design, and data management. M.M. and H.M. revised the manuscript for intellectual content. Y.Y. and T.A. provided overall supervision. All the authors have read and approved the final version of the manuscript.

## Nonstandard Abbreviations and Acronyms

CSVD: cerebral small vessel disease
DWMLs: deep white matter lesions
eGFR: estimated glomerular filtration rate
Na/K ratio: sodium-to-potassium ratio

## References

1. Fazekas F, Kleinert R, Offenbacher H, Schmidt R, Kleinert G, Payer F, Radner H, Lechner H. Pathologic correlates of incidental MRI white matter signal hyperintensities. Neurology. 1993;43:1683–1689. doi: 10.1212/wnl.43.9.1683.

2. Wardlaw JM, Smith EE, Biessels GJ, Cordonnier C, Fazekas F, Frayne R, Lindley RI, O’Brien JT, Barkhof F, Benavente OR, et al. Neuroimaging standards for research into small vessel disease and its contribution to ageing and neurodegeneration. Lancet Neurol. 2013;12:822–838. doi: 10.1016/S1474-4422(13)70124-8.

3. Debette S, Markus HS. The clinical importance of white matter hyperintensities on brain magnetic resonance imaging: systematic review and meta-analysis. BMJ. 2010;341:c3666. doi: 10.1136/bmj.c3666.

4. Pantoni L. Cerebral small vessel disease: from pathogenesis and clinical characteristics to therapeutic challenges. Lancet Neurol. 2010;9:689–701. doi:10.1016/S1474-4422(10)70104-6.

5. Kawasaki T, Itoh K, Uezono K, Sasaki H. A simple method for estimating 24 h urinary sodium and potassium excretion from second morning voiding urine specimen in adults. Clin Exp Pharmacol Physiol. 1993;20:7–14. doi: 10.1111/j.1440-1681.1993.tb01496.x.

6. O’Donnell M, Mente A, Rangarajan S, McQueen MJ, Wang X, Liu L, Yan H, Lee SF, Mony P, Devanath A, et al. Urinary sodium and potassium excretion, mortality, and cardiovascular events. N Engl J Med. 2014;371:612–623. doi: 10.1056/NEJMoa1311889.

7. Cogswell ME, Loria CM, Terry AL, Zhao L, Wang CY, Chen TC, Wright JD, Pfeiffer CM, Merritt R, Moy CS, et al. Estimated 24-hour urinary sodium and potassium excretion in US adults. JAMA. 2018;319:1209–1220. doi: 10.1001/jama.2018.1156.

8. World Health Organization. Guideline: sodium intake for adults and children. Geneva: World Health Organization; 2012.

9. Hisamatsu T, Kogure M, Tabara Y, Hozawa A, Sakima A, Tsuchihashi T, Yoshita K, Hayabuchi H, Node K, Takemi Y, et al. Practical use and target value of urine sodium-to-potassium ratio in assessment of hypertension risk for Japanese: Consensus Statement by the Japanese Society of Hypertension Working Group on Urine Sodium-to-Potassium Ratio. Hypertens Res. 2024;47:3288–3302. doi: 10.1038/s41440-024-01861-x.

10. Ito S, Nagasawa T, Abe M, Mori T. Strain vessel hypothesis: a viewpoint for linkage of albuminuria and cerebro-cardiovascular risk. Hypertens Res. 2009;32(2):115–121. doi: 10.1038/hr.2008.27.

11. Georgakis MK, Chatzopoulou D, Tsivgoulis G, Petridou ET. Albuminuria and cerebral small vessel disease: a systematic review and meta-analysis. J Am Geriatr Soc. 2018;66:509–517. doi: 10.1111/jgs.15240.

12. Larsson SC, Orsini N, Wolk A. Dietary potassium intake and risk of stroke: a dose–response meta-analysis of prospective studies. Stroke. 2011;42(10):2746–2750. doi:10.1161/STROKEAHA.111.622142.

13. Vinceti M, Filippini T, Crippa A, de Sesmaisons-Lecarré A, Wise LA, Orsini N. Meta-analysis of potassium intake and the risk of stroke. J Am Heart Assoc. 2016;5:e004210. doi:10.1161/JAHA.116.004210.

14. O’Donnell M, Yusuf S, Vogt L, Mente A, Messerli FH. Potassium intake: the Cinderella electrolyte. Eur Heart J. 2023;44:4925–4934. doi:10.1093/eurheartj/ehad628.

15. D’Elia L, Cappuccio FP, Masulli M, La Fata E, Rendina D, Galletti F. Effect of potassium supplementation on endothelial function: a systematic review and meta-analysis of intervention studies. Nutrients. 2023;15:853. doi:10.3390/nu15040853.

16. Iwahori T, Miura K, Ueshima H. Time to consider use of the sodium-to-potassium ratio for practical sodium reduction and potassium increase. Nutrients. 2017;9(7):700. doi:10.3390/nu9070700.

17. Perez V, Chang ET. Sodium-to-potassium ratio and blood pressure, hypertension, and related factors. Adv Nutr. 2014;5:712–741. doi:10.3945/an.114.006783.

18. Zanetti D, Bergman H, Burgess S, Assimes TL, Bhalla V, Ingelsson E. Urinary albumin, sodium, and potassium and cardiovascular outcomes in the UK Biobank: observational and Mendelian randomization analyses. Hypertension. 2020;75(3):714–722. doi:10.1161/HYPERTENSIONAHA.119.14028.

19. Yamasaki K, Hata J, Furuta Y, Hirabayashi N, Ohara T, Yoshida D, Hirakawa Y, Nakano T, Kitazono T, Ninomiya T. Association of albuminuria with white matter hyperintensities volume on brain magnetic resonance imaging in elderly Japanese – the Hisayama study. Circ J. 2020;84:935–942. doi:10.1253/circj.CJ-19-1069.

20. Cunha MR, Cunha AR, Marques BCAA, Mattos SS, D’El-Rei J, França NM, Oigman W, Neves MF. Association of urinary sodium/potassium ratio with structural and functional vascular changes in non-diabetic hypertensive patients. J Clin Hypertens (Greenwich). 2019;21:1360–1369. doi:10.1111/jch.13660.

21. Gansevoort RT, Lambers Heerspink HJ, Witte EC. Methodology of screening for albuminuria. Nephrol Dial Transplant. 2007;22:2109–2111. doi:10.1093/ndt/gfm267.

22. McLean RM. Measuring population sodium intake: a review of methods. Nutrients. 2014;6:4651–4662. doi:10.3390/nu6114651.

23. Joo L, Shim WH, Suh CH, Lim SJ, Heo H, Kim WS, Hong E, Lee D, Sung J, Lim JS, et al. Diagnostic performance of deep learning-based automatic white matter hyperintensity segmentation for classification of the Fazekas scale and differentiation of subcortical vascular dementia. PLoS One. 2022;17(9):e0274562. doi:10.1371/journal.pone.0274562.

